# A Novel Explainable AI Method to Assess Associations between Temporal Patterns in Patient Trajectories and Adverse Outcome Risks: Analyzing Fitness as a Risk Factor of ADRD

**DOI:** 10.1101/2024.05.17.24307541

**Authors:** Yijun Shao, Edward Y. Zamrini, Ali Ahmed, Yan Cheng, Stuart J. Nelson, Peter Kokkinos, Qing Zeng-Treitler

## Abstract

We present a novel explainable artificial intelligence (XAI) method to assess the associations between the temporal patterns in the patient trajectories recorded in longitudinal clinical data and the adverse outcome risks, through explanations for a type of deep neural network model called Hybrid Value-Aware Transformer (HVAT) model. The HVAT models can learn jointly from longitudinal and non-longitudinal clinical data, and in particular can leverage the time-varying numerical values associated with the clinical codes or concepts within the longitudinal data for outcome prediction. The key component of the XAI method is the definitions of two derived variables, the temporal mean and the temporal slope, which are defined for the clinical concepts with associated time-varying numerical values. The two variables represent the overall level and the rate of change over time, respectively, in the trajectory formed by the values associated with the clinical concept. Two operations on the original values are designed for changing the values of the two derived variables separately. The effects of the two variables on the outcome risks learned by the HVAT model are calculated in terms of impact scores and impacts. Interpretations of the impact scores and impacts as being similar to those of odds ratios are also provided. We applied the XAI method to the study of cardiorespiratory fitness (CRF) as a risk factor of Alzheimer’s disease and related dementias (ADRD). Using a retrospective case-control study design, we found that each one-unit increase in the overall CRF level is associated with a 5% reduction in ADRD risk, while each one-unit increase in the changing rate of CRF over time is associated with a 1% reduction. A closer investigation revealed that the association between the changing rate of CRF level and the ADRD risk is nonlinear, or more specifically, approximately piecewise linear along the axis of the changing rate on two pieces: the piece of negative changing rates and the piece of positive changing rates.

## 1. Introduction

The emergence of deep learning techniques with deep neural networks (DNNs) as the main tools has brought remarkable advancements to the field of artificial intelligence (AI) over the past decade.^1-6^ One significant challenge posed by the DNN models is their inherent complexity, making them opaque and difficult to understand for humans - a characteristic commonly referred to as the “black-box” nature of these models.^7,8^ Explainable AI (XAI) techniques aim to unravel the decision-making process of the black-box models.^9,10^ By offering interpretability and increasing transparency, XAI can bridge the gap between DNN models and human comprehension, empowering users to understand, validate, and trust the outcomes of the powerful AI systems.^11,12^ One class of the existing XAI methods provide explanations by quantifying the influence of the features or variables on the model predictions. Those methods include Local Interpretable Model-agnostic Explanations (LIME),^13^ SHapley Additive exPlanations (SHAP),^14,15^ Layer-wise Relevance Propagation (LRP),^16^ and feature importance.^17,18^

In contrast to DNN models, conventional statistical models, when viewed as machine learning model, are considered explainable, because all their inner parameters are readily interpreted.^18^ For example, for logistic regression models, the influence of the variables on the prediction is explicitly quantified by the corresponding coefficients in the model and is interpreted as log-odds ratios. Motivated partially by the explanations for logistic regression models, we previously developed an XAI method for the type of DNN models that take tabular data as inputs. This method produces impact scores for the input variables, which have similar interpretations as the coefficients in logistic regression models.^19,20^ Therefore, the impact scores can be used for risk factor analysis in a way similar to the coefficients in logistic regression models.

Transformer, a new type of DNN architecture developed recently,^21^ has revolutionized the field of natural language processing (NLP), as demonstrated by the development of the large language models such as Bidirectional Encoder Representations from Transformers (BERT),^22^ Generative Pre-trained Transformer (GPT),^23-25^ and Large Language Model Meta AI (LLaMa).^26^ Encouraged by this great success and based on the similarities between longitudinal clinical data and natural language data, we designed a Transformer-based architecture, referred to as Hybrid Value-Aware Transformer (HVAT),^27^ for jointly learning from longitudinal clinical data and non-longitudinal data. In particular, HVAT can leverage the time-varying numerical values associated with the clinical codes/concepts in the longitudinal data for outcome prediction. Examples of such clinical concepts include lab test orders with associated lab values, vital signs with associated vital sign values. It has been demonstrated that HVAT models can achieve excellent predictive performance and outperform their counterparts with the same input data except that the longitudinal data is converted to non-longitudinal data (e.g., through aggregation over time).^27^ While the longitudinal data are records of the patient trajectories, which often carry various temporal patterns, such information would be lost after the conversion to non-longitudinal data. We believe HVAT models can recognize those temporal patterns and leverage such information in the learning process for better performance. The questions are: Can we calculate how the temporal patterns impact the predictions in the HVAT models? If so, can we also make the calculation results clinically understandable?

To answer these questions, we have designed a novel XAI method, which makes the HVAT models explainable by quantifying the associations between the temporal patterns in the patient trajectories and the predicted adverse outcome risks. As an initial attempt, we are focused on the temporal patterns in the clinical concepts with associated numerical values. We demonstrate the use of the method by applying it to the study of cardiorespiratory fitness (CRF) as a risk factor of Alzheimer’s disease and related dementias (ADRD). Patient CRF is commonly measured using exercise tolerance testing and expressed as metabolic equivalents (METs) with larger values representing higher CRF levels.^28^ The XAI method is used to assess the associations between the temporal patterns in CRF levels and the risks of ADRD as the adverse outcome.

## 2. Methods

In this section, we describe the XAI method as a general framework, so that it is not limited to the analysis of CRF in this study but can also be used to analyze other various clinical variables.

### 2.1 HVAT architecture

Since the HVAT architecture has been previously described in full detail,^27^ we will only briefly describe the part that is relevant to the XAI method here.

To use HVAT for modeling, the first step is to format the longitudinal data in a special way as follows. For each patient, a time window is specified on the history data, and the endpoint of the window is called the index time/date. The time window is divided into smaller equal-length intervals, which are then indexed by the natural numbers: 1, 2, 3, …, backwards in time (i.e., from the end to the start). The natural numbers are called the temporal indices. The time window may vary by patient, but the length of the small intervals is the same for all patients. Next, the clinical data within the time window for the patient is represented as a sequence of clinical tokens. A clinical token is a triple (*t, C, v*), where *t* is a temporal index, *C* is a clinical concept, and *v* is either a numerical value associated with *C* or the default value zero. The presence of a clinical token (*t, C, v*) in the sequence means that the clinical concept *C* occurred one or more times within the time interval with temporal index *t*. If *C* is a clinical concept with associated values, the *v* is the value of *C* if *C* only occurred once, or an aggregation of the multiple values if *C* occurred multiple times. The aggregation method may be different for different concepts. If *C* is a clinical concept without values (e.g., a diagnosis), then *v* is set as the default value zero. The HVAT model takes the sequence of clinical tokens as part of the input data (the other part is the non-longitudinal data in tabular format), and outputs a single value *p* between 0 and 1 representing the probability of the adverse outcome.

### 2.2 Temporal mean and temporal slope

A clinical concept *C* with associated numerical values is effectively a temporal variable, and the multiple values at multiple temporal indices effectively form a time series. To reveal how the temporal patterns of the time series are associated with the predicted risks, we consider the two temporal patterns characterized by, respectively, the two variables: the temporal mean *α* and the temporal slope *β*. The two variables are defined as follows. Suppose on a patient the temporal variable *C* takes values *v*_1_, …, *v*_*n*_ at temporal indices *t*_1_, …, *t*_*n*_, respectively (*n* may vary over the patients). Then the *temporal mean α* is defined simply as the mean of the values:

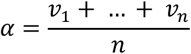

It represents the *overall level* of *C* on the patient during the chosen time window. The *temporal slope β* is defined as the slope of the line fitted to the data points (−*t*_1_, *v*_1_), …, (−*t*_*n*_, *v*_*n*_) in the time-value plane using a linear regression. Note that the temporal indices are ordered backwards on the time axis, and adding the minus signs makes the time components of the data points ordered forwardly along the time axis. Let 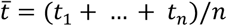 and let (−*t, v*) represent an arbitrary point on the line, then the equation of the fitted line is 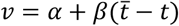. The slope *β* can be calculated explicitly using the formula^29^

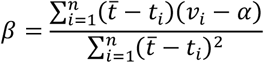

It characterizes the *linear trend* of *C* on the individual patient. In particular, *β* > 0 means a linearly increasing trend over time, *β* < 0 means a linearly decreasing trend over time, and *β* = 0 means the trend of staying at the same level over time. Note that a linear regression requires at least two data points, therefore the slope *β* is only defined when *n* ≥ 2. In contrast, the mean *α* is defined for all *n* ≥ 1. The two variables *α* and *β* are *derived variables*, since they are not among the original variables but are derived from them.

The two derived variables *α* and *β* are “orthogonal” to each other, meaning that we can change the value of one of them while keeping the other value unchanged by changing the original sequence of values in some special way. This property is important because it is necessary to separate the effects of the two derived variables on the outcome. Specifically, given a sequence of values *v*_1_, …, *v*_*n*_ at temporal indices *t*_1_, …, *t*_*n*_, respectively, with temporal mean *α* = *α*_0_ and the temporal slope *β* = *β*_0_, to change the time series {*v*_*i*_} to a new value time series 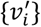 of the same length, we can perform one of the following two operations (see Figure 1 for illustration):

**Figure 1.**
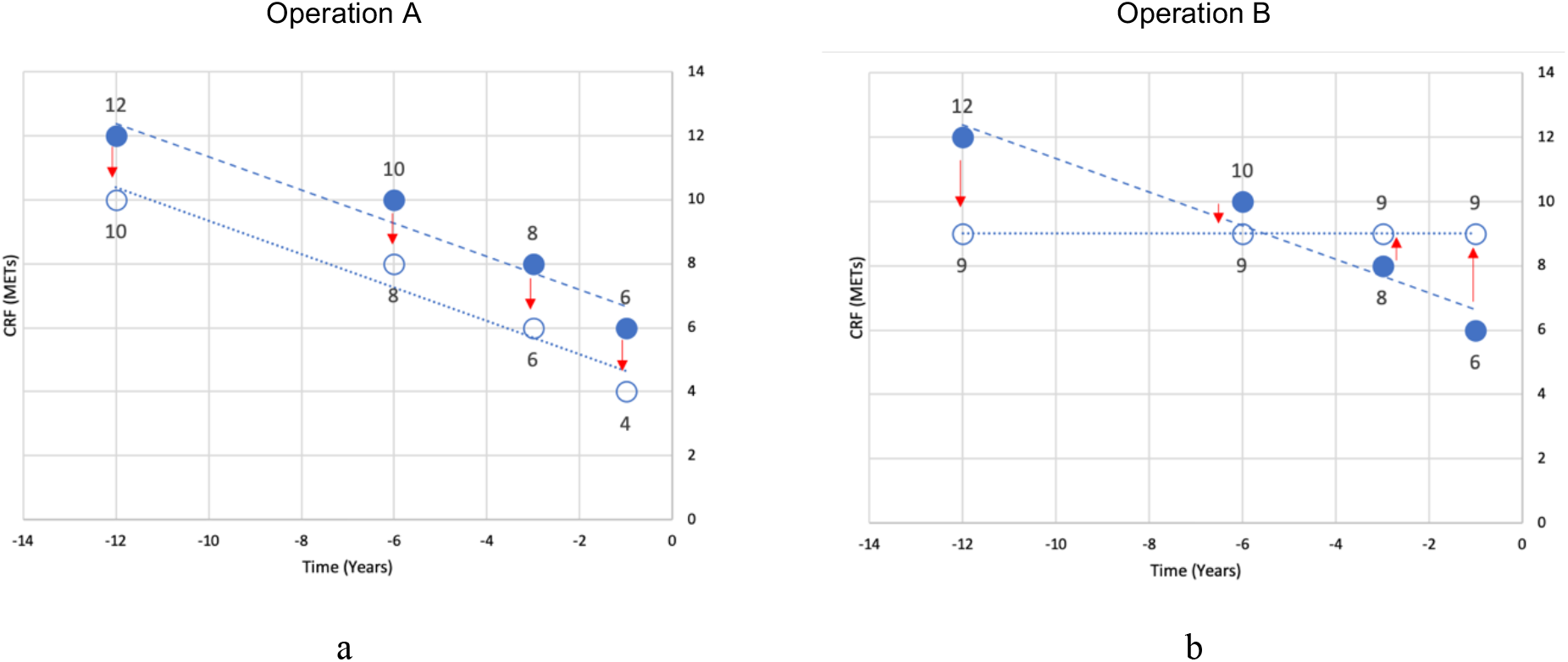
An illustration of Operations A and B through a concrete example. The 4 solid dots in (a) and (b) represent the 4 CRF values (12, 10, 8, 6) of a patient at the temporal indices (12, 6, 3, 1). In (a), Operation A shifts the 4 dots down by 2 METs to the 4 circles, so that the original temporal mean 9.0 is lowered to the reference value 7.0, while the temporal slope is maintained. In (b), Operation B moves the 4 dots to the 4 circles on a horizontal line at level 9.0, so that the temporal slope is changed from originally a negative number to the reference value 0, while the temporal mean (9.0) is maintained.

- Operation A: Given a new value *α*_1_ of the temporal mean, set 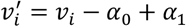.
- Operation B: Given a new value *β*_1_ of the temporal slope, set 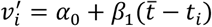.

It is easy to verify that Operation A changes the temporal mean from *α*_0_ to *α*_1_ but keeps the temporal slope at *β*_0_, while Operation B changes the temporal slope from *β*_0_ to *β*_1_ but keeps the temporal mean at *α*_0_.

Next, we define the impact and the impact score for each of the two derived variables. Suppose we have a well-trained HVAT model and suppose also that the risk scores output by the model can be interpreted as the probability of having the adverse outcome. The latter can be empirically verified by plotting the calibration curve: if the calibration curve approximates the diagonal line well, then it means the risk scores are good for use. Otherwise, we need to recalibrate the risk scores and use the new scores as the output of the HVAT model.

### 2.3 The impact and the impact score of the temporal mean

First, calculate the temporal mean values on all patients in the cohort. Then choose and fix a value as the baseline for the temporal mean, which will be called the reference value and denoted as *α*_*r*_. Let log*i*t 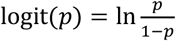 denote the logit function, which is a bijective mapping from the interval (0, 1) to the entire set of real numbers (−∞, +∞). Next, for a patient with the temporal mean value *α*_*c*_ ≠ *α*_*r*_, take the following steps:

1. Evaluate the model using the input data of the patient. Let *p*_+_ denote the model output.
2. Change the patient’s time series data using Operation A with *α*_1_ ≠ *α*_*r*_, and re-evaluate the model using the modified input data. Let *p*_(_ denote the model output.
3. The (*individual-level*) *impact of the temporal mean on this patient* is defined as the difference log*i*t(*p*_*c*_) − log*i*t(*p*_*r*_).
4. The (*individual-level*) *impact score of the temporal mean on this patient* is defined as the difference ratio

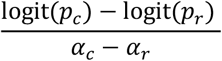

### 2.4 The impact and the impact score of the temporal slope

First, calculate the temporal slopes for all patients in the cohort who have at least two values at two different temporal indices. Then choose and fix a value as the baseline for the temporal slope, which will be called the reference value and denoted as *β*_*r*_.

Next, for a patient with the temporal slope value *β*_+_ ≠ *β*_(_, take the following steps:

1. Evaluate the model using the input data of the patient. Let *p*_*c*_ denote the model output.
2. Change the patient’s value sequence to a new value sequence using Operation B with *β*_1_ = *β*_*r*_, and re-evaluate the model using the modified input data. Let *p*_*r*_ denote the model output.
3. The (*individual-level*) *impact of the temporal slope on this patient* is defined as the difference log*i*t(*p*_*c*_) − log*i*t(*p*_*r*_).
4. The (*individual-level*) *impact score of the temporal slope on this patient* is defined as the difference ratio

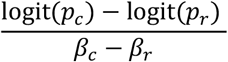

### 2.5 The impact and the impact score at (sub)population level

Given a subpopulation, the mean of the impact (resp. the mean of the impact scores) on all the patients in the subpopulation (excluding those which are undefined) is defined as the subpopulation-level impact (resp. subpopulation-level impact score). When the subpopulation is the entire cohort, this defines the population-level impact (resp. population-level impact score), which will often simply be called the variable’s impact (resp. impact score).

### 2.6 Impact score by value and impact by value

For each value *x*_*i*_ of a (derived) variable *x*, we define the *impact score of* the variable *at the value x*_*i*_ as the mean of the individual-level impact scores on all those patients with value *x*_*i*_. This is a special case of the subpopulation-level impact scores. We also define the *impact of* the variable *at the value x*_*i*_ as the mean of the individual-level impact on all the patients with value *x*_*i*_.

The impact score (respectively (resp.) impact) by value is effectively a function which describes how the impact score (resp. impact) changes along with the variable *x*, and hence provides much more information than the single-valued population-level impact score (resp. impact). One motivation for the definitions is that one can tell how nonlinear (or how far away from being linear) the association between the variable and the outcome is from the functions. If the association is linear, then the impact score (resp. impact) by value should be a constant (resp. linear) function of *x*. The more the impact score (resp. impact) is far away from being constant (resp. linear), the more nonlinear the association is. Graphically, the impact score (resp. impact) by value can be visualized as a curve, and the closer to being a straight horizontal line (resp. straight line) the curve is, the closer to being linear the association is. Moreover, if the association is linear, then the slope of the line for the impact by value is exactly the y-value of the horizontal line for the impact score by value.

The definition of the impact is theoretically valid but practically infeasible to calculate, because a continuous variable can take infinitely many values, while practically we only have in the dataset a sample of finitely many patients, who contribute only finitely many values. Therefore, away from those finitely many values, there are still infinitely many values which no patients take, and the impact score at those values are literally undefined by the above definition. This situation is similar to finding the distribution (in terms of probability density function) of a continuous random variable using a finite sample. In that case, the solution is approximation by a histogram, where the distribution is estimated on a finite number of bins with each bin containing sufficiently many data points. That inspires us to take the following approach to estimate the impact/impact score by value.

First, we determine a value range expressed as an interval for the values of the (derived) variable. The range can be smaller than the theoretically largest possible range. Next, we divide the range into small bins, which can have different widths, and choose and fix a value from each bin as the representative value for that bin. For each bin, we calculate the mean of the individual-level impact (resp. impact score) over the bin, and regard it as the impact (resp. impact score) of the value representing the bin. For stability of the mean values, every bin should contain sufficiently many (e.g., >=100) patients. Then by linear extrapolation we obtain the impact (resp. impact score) for all values over the range. Graphically, it is equivalent to plot a curve by straightly connecting the dots which represent the mean impact/impact scores for the representative values of the bins.

### 2.7 Interpretations

The impact and impact score are closely related to odds ratios. Actually, the individual-level impact for a patient is the (natural) log of an odds ratio:

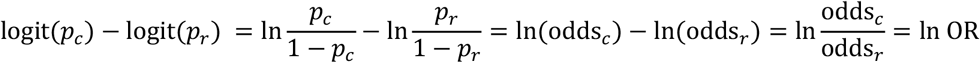

where 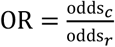 is the odds ratio. Therefore, the odds ratio OR can be obtained by simply exponentiating the impact: OR = exp (individual-level impact). Note that this shows the importance of ensuring the model has good calibration, because for the term 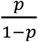 to be interpreted as an odds, the *p* must be interpreted as a probability.

For the impact by value, which is a subpopulation-level impact, we can also define the odds ratio as OR = exp (impact by value), and it is easy to see that this OR is the geometric mean of the individual odds ratios over that subpopulation.

Suppose a variable *x* takes value *x*_*c*_ on the patient and the reference value is *x*_*r*_. Let | · | denote the absolute value. Then we can interpret the individual-level odds ratio calculated on a patient as follows:

- Compared to the hypothetical situation that the variable *x* takes value *x*_*r*_, the actual situation of *x* taking value *x*_*c*_ increases (if OR>1; or reduces if OR<1) the odds of having the adverse outcome by |OR − 1| × 100% on the patient.

For the impact by value, i.e., the mean impact of a value *x*_*i*_, the odds ratio is OR = exp (impact by value). We can interpret the odds ratio as follows:

- Compared to the situation that the variable *x* takes value *x*_*r*_, the variable *x* taking value *x*_*i*_ increases (if OR>1; or reduces if OR<1) the odds of having the adverse outcome by |OR − 1| × 100% on average.

As the individual-level impact score is a normalized individual-level impact, exponentiating it also gives an odds ratio: OR = exp(individual-level impact score). The difference is that the comparison is between before and after a one-unit change in the variable. We can interpret this odds ratio as follows:

- Each one-unit change in the variable is, on average, associated with an increase (if OR>1; or reduction if OR<1) in the odds of having the (derived) adverse outcome by |OR − 1| × 100% on the patient.

The subpopulation-level impact score is a normalized subpopulation-level impact, and the odds ratio can be defined as OR = exp(subpopulation-level impact score). This applies to in particular the impact score by value. We can interpret the odds ratio as follows:

- Each one-unit change in the (derived) variable is, on average, associated with an increase (if OR>1; or reduction if OR<1) in odds of having the adverse outcome by |OR − 1| × 100%.

## 3. Experiment

### 3.1 Dataset

We performed a retrospective case-control study using the U.S. Veterans’ electronic health records data administered by the U.S. Department of Veterans Affairs. Using a NLP tool developed by our team earlier,^30^ we identified 538 thousand patients who had at least one CRF value in METs documented in the textual notes. From them, we further identified 51 thousand patients who received an ADRD diagnosis on or before 12/31/2019, from which we randomly selected 50,000. This was the case group. For the control group, we randomly selected 50,000 from those who had never received an ADRD diagnosis and were still alive on 12/31/2019. The final cohort was the combination of the case group and the control group.

We defined the *index date* for each case as the date one year before the first ADRD diagnosis and for each control the fixed date 12/31/2018 (one year before 12/31/2019). The one-year gap was to ensure that the cases were free of ADRD before the index date and also ensure the controls were free of ADRD one year after the index date.

### 3.2 Model development

A HVAT model was trained to classify the patients by their ADRD status at one year after the index date using the EHR data up to the index date. The outcome variable was naturally a binary variable, which was coded as: 1 = case, 0 = control. The input data was the EHR data recorded within the time window from 1/1/2000 to the index date. The time window was divided into intervals of one year long for temporal index assignment. The longitudinal data only included CRF values expressed in METs, which means we had only one clinical concept, i.e., the CRF. When there were multiple CRF values over the same time interval, we used the maximum as the aggregation method for that time interval. CRF was treated as a continuous variable, and their associated values were all rounded to the first decimal place. In this study, we restricted the range of CRF values to be between 2.0 METs and 23.9 METs. The non-longitudinal data included age (at one year after index date), sex, race, and ethnicity. For the model, we set embedding dimension *d* = 32 and number of Transformer blocks *N* = 2.

The cohort was randomly split into 3 sets: training (80%), validation (10%) and testing (10%). The training process was stopped when the performance on the validation set plateaued. The performance on the testing set was reported as the final model performance. The primary metric of model performance was the area under ROC curve (AUC). A threshold on the output scores to optimize the classification accuracy was chosen, and then accuracy, sensitivity and specificity were evaluated based on the threshold.

### 3.3 Application of XAI

We calculated the impacts and the impact scores for the temporal mean *α* and the temporal slope *β* of the CRF variable. For the temporal mean, we used the median value over the population as the reference value, which was 7.0, and for the temporal slope, we also used the median value over the population as the reference value, which was 0.0. Below we illustrate the XAI method, especially Operations A and B, through a concrete example, as in Figure 1.

To calculate the impact and impact score by value for the temporal mean of CRF, we set the range of temporal mean to be [2, 19] (the range of the raw data was [2, 24]), and divided the range into bins: [2, 2.5), [2.5, 3.5), [3.5, 4.5), …, [18.5, 19], all having a width of 1 except for the first and the last which had a width of 0.5. The representative values for the bins were set as the integers 2, 3, 4, …, 19.

To calculate the impact and impact score by value for the temporal slope of CRF (measured in METs per decade), we set the range of the temporal slope to be [-6, 6] because an increase or decrease by more than 6 METs per decade over several years was rarely observed. Then we divided this range into 12 small bins [-6, -5), [-5, -4), …, [5, 6], all having a width of 1. The representative values for the bins were set as -5.5, -4.5, …, 4.5, and 5.5.

## 4. Results

A summary of the demographic and clinical characteristics for cases and controls is shown in Table 1.

**Table 1.** Demographic and clinical characteristics for cases and controls.

|  | Cases (N=50,000) | Controls (N=50,000) |
| --- | --- | --- |
| Age at Endpoint |  |  |
| Mean ± SD | 76.4 ± 9.5 | 69.2 ± 10.1 |
| Median (Q1, Q3) | 77.1 (69.9, 83.6) | 70.7 (63.2, 74.8) |
| Sex |  |  |
| Female | 1,032 (2.1%) | 3,360 (6.7%) |
| Male | 48,968 (97.9%) | 46,640 (93.3%) |
| Race |  |  |
| Black | 7,387 (14.8%) | 8,682 (17.4%) |
| White | 38,408 (76.8%) | 36,820 (73.6%) |
| Other | 1,030 (2.1%) | 1,327 (2.7%) |
| Unknown | 3,175 (6.3%) | 3,171 (6.3%) |
| Ethnicity |  |  |
| Hispanic | 2,983 (6.0%) | 2,867 (5.7%) |
| Non-Hispanic | 44,695 (89.4%) | 44,861 (89.7%) |
| Unknown | 2,322 (4.6%) | 2,272 (4.5%) |
| CRF - Temporal mean |  |  |
| Mean ± SD | 6.9 ± 2.7 | 8.4 ± 3.0 |
| Median (Q1, Q3) | 7.0 (4.7, 8.7) | 8.3 (6.6, 10.2) |
| CRF - Temporal slope |  |  |
| Mean ± SD | -0.030 ± 0.491 | -0.025 ± 0.488 |
| Median (Q1, Q3) | 0.0 (0.0, 0.0) | 0.0 (0.0, 0.0) |

A HVAT model was trained on the training set, and its classification performance on the testing set was: AUC = 0.81, Sensitivity = 0.70, Specificity = 0.75, Accuracy = 0.73.

Figure 2 shows the calibration curve of the model. We can see that the curve fits to the diagonal line well, which confirms that the predicted risk scores well represent the empirical probabilities of having ADRD. Therefore, we can calculate the impact scores and impacts using the predicted risk scores directly without any further calibration.

**Figure 2.**
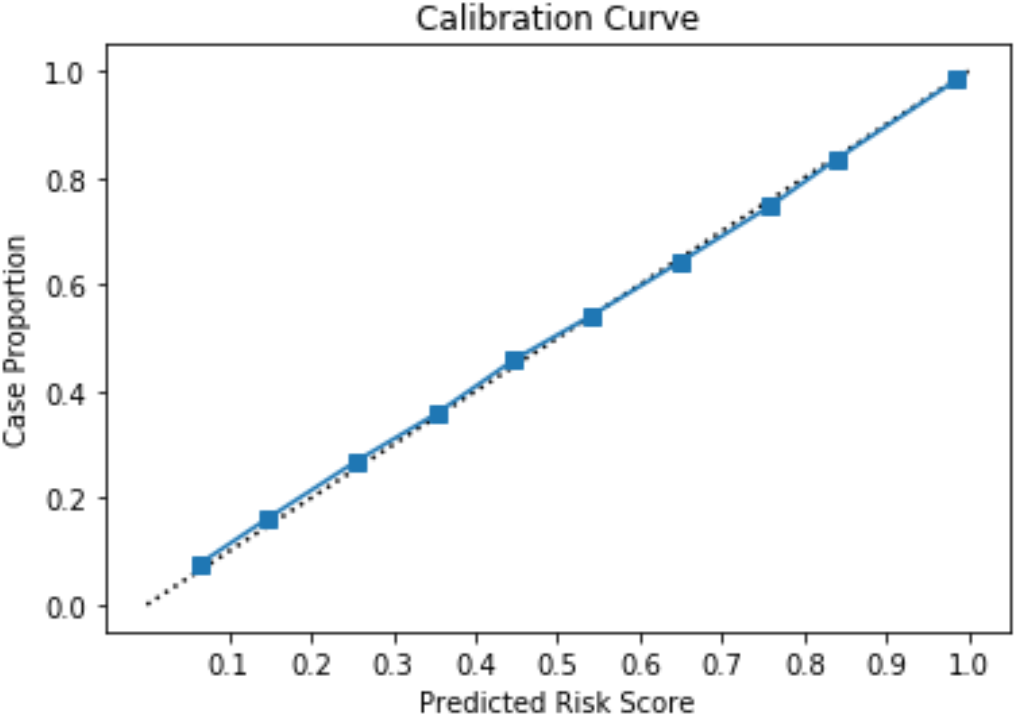
The calibration curve of the HVAT model.

For *α* the temporal mean of CRF, the population-level impact score was -0.052 per MET, and the corresponding odds ratio was OR = exp (−0.052) = 0.949. The interpretation of this result is:

Each one MET increase in temporal mean of CRF is, on average, associated with a reduction by (1-0.949) x100% = 5.1% in the odds of having ADRD, assuming all the demographic characteristics and the temporal slope of CRF are held the same.

The impact score by value and impact by value are shown in Figure 3 as blue curves. The individual-level impacts and impact scores are also shown in Figure 3 as orange dots. The straight dashed purple line in Figure 3a represents the population-level impact score.

**Figure 3.**
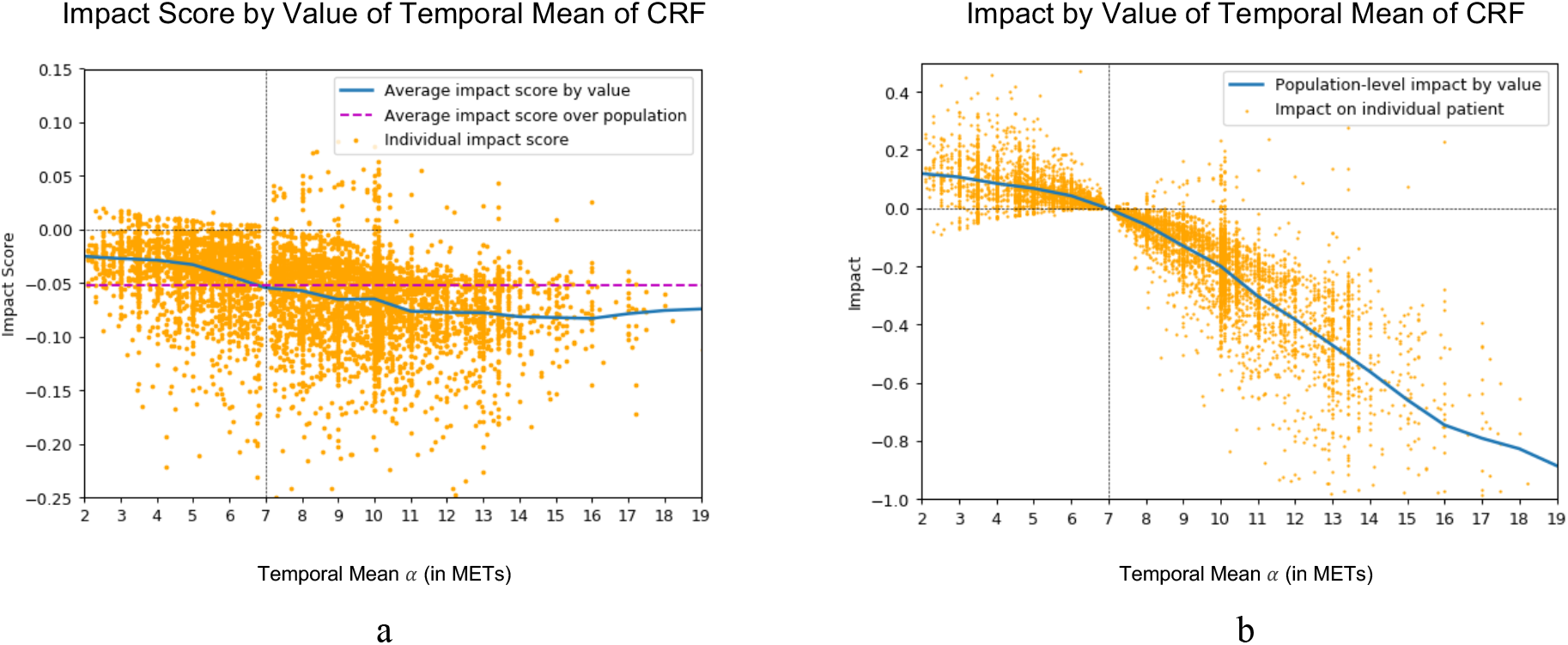
The impact score by value and the impact by value of the temporal mean of CRF.

In Figure 3a, the blue curve shows how the impact score by value of the temporal mean of CRF (y-value) changes along with the temporal mean of CRF (x-value). For each x-value, the y-value of the point on the blue curve of the x-value is an average of all the y-values of the orange dots having approximately the same x-values. We can see that the impact score by value is negative over the entire range, which means that, on average, at each value of temporal mean, an increase in that value is associated with a reduction in the ADRD risk. However, the amount of reduction varies slightly with the x-values: the reduction is smaller for patients with lower temporal means and larger for those with higher temporal means.

In Figure 3b, the blue curve represents how the impact of the temporal mean of CRF (y-value) changes with the temporal mean of CRF (x-value). Like Figure 3a, this curve is also a value-wise average of the individual impacts. We see that the curve is decreasing over the entire range, which means that, on average, an increase in temporal mean of CRF is associated with a reduction in ADRD risk. However, the slope of curve varies slightly with the x-values: the curve is flatter for lower temporal means while steeper for higher temporal means, which means the reduction in ADRD risk is smaller for lower temporal means and larger for higher temporal means.

For *β* the temporal slope of CRF, the population-level impact score was -0.0106 per MET/decade, and the odds ratio was OR = exp (−0.0106) = 0.989. This result was interpreted as follows:

Each one unit increase in the temporal slope of CRF is, on average, associated with a reduction by (1-0.989) x100% = 1.1% in the odds of having ADRD, assuming all the demographic characteristics and the temporal mean of CRF are held the same.

The impact score by value and the impact by value of the temporal slope are shown in Figure 4 as blue curves, along with the individual-level versions as orange dots.

**Figure 4.**
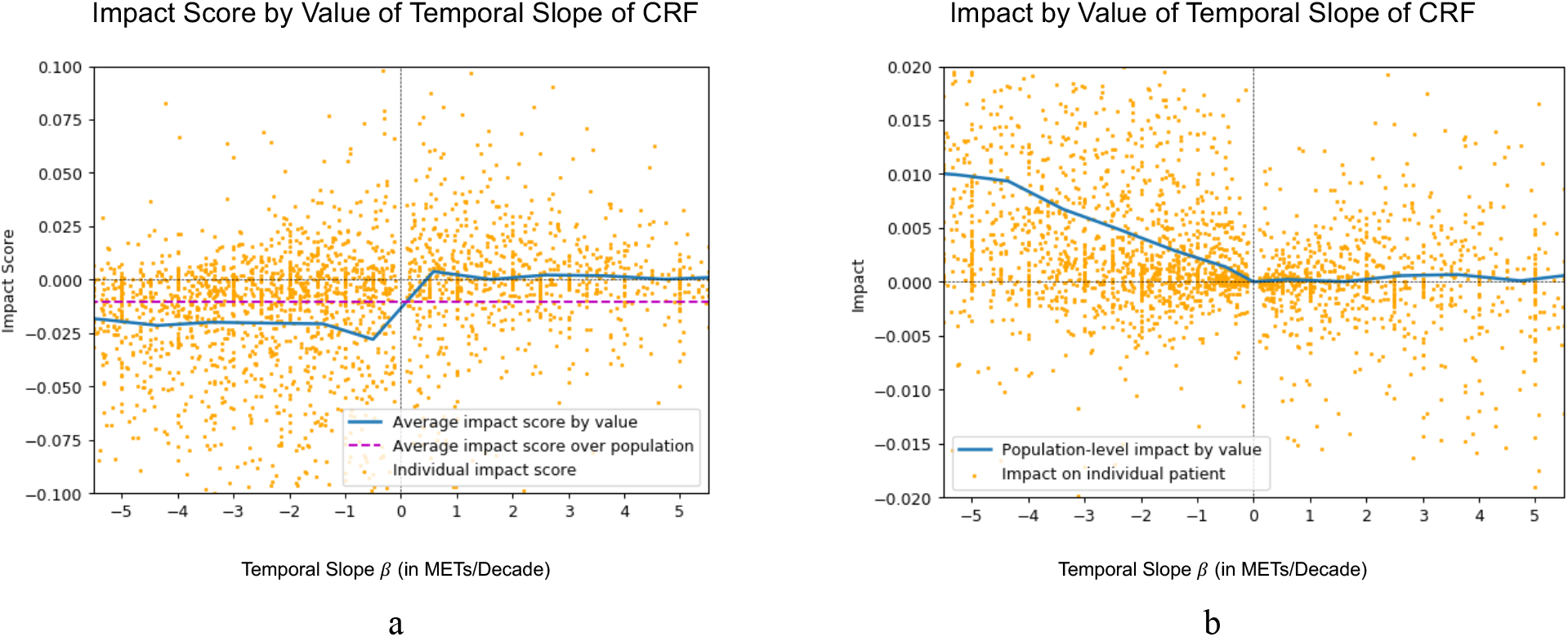
The impact score by value and the impact by value of the temporal slope of CRF.

In Figure 4a, the blue curve shows how the impact score by value of the temporal slope of CRF (y-value) changes with the temporal slope of CRF (x-value). We can see that the impact score is negative and stays at about the same level for the negative temporal slopes (i.e., *β* < 0), and the impact score is approximately zero for the positive temporal slopes (i.e., *β* > 0). There is an abrupt increase (or a jump) of the impact score from a negative value to zero from the left side to the right side of *β* = 0. This phenomenon is more clearly shown in Figure 4b.

In Figure 4b, the blue curve shows how the impact by value of the temporal slope of CRF (y-value) changes with the temporal slope of CRF (x-value). We can see that the curve represents approximately a piece-wise linear function: the curve is approximately a straight line with a negative slope for *β* < 0, and approximately a horizontal line (zero slope) for *β* > 0. The abrupt change of the slope of the blue curve from negative to zero occurs at the point *β* = 0. (Note that the slope of the curve should not be confused with the temporal slope.) This change matches with the abrupt change from negative impact score to zero impact score in Figure 4a.

This shows that, the subpopulation with a decreasing trend of CRF (i.e., *β* < 0) and that with an increasing trend of CRF (i.e., *β* > 0) have distinct characteristics in terms of the impact scores of *β*, while within each subpopulation, the patients are similar.

This observation motivated us to calculate two subpopulation-level impact scores of *β*: the impact score of *β* = -0.018 (per MET/decade) on the subpopulation with *β* < 0, and the impact score of *β* = 0.002 (per MET/decade) on the subpopulation with *β* > 0. So, we see much stronger effect of the temporal slope on the subpopulation with *β* < 0 than on the entire population, while near zero effect of the temporal slope on the subpopulation with *β* > 0.

The interpretations for the two subpopulation-level impact scores are:

- For patients with a decreasing trend in CRF, each one-unit reduction in the speed of CRF decreasing is, on average, associated with a reduction by (1-exp (−0.018)) x 100% = 1.8% in the odds of having ADRD.
- For patients with an increasing trend in CRF, each one-unit increase in the speed of CRF increasing is, on average, associated with almost no change in the odds of having ADRD.

## 5. Discussion and Conclusions

In this study, we developed a new XAI method for explaining a type of Transformer-based DNN models referred to as HVAT models. These models can learn from longitudinal and non-longitudinal data jointly, and in particular can leverage the time-varying numerical values associated with the clinical codes or concepts in the longitudinal data. The XAI method is focused on quantifying the associations between the linear temporal patterns in the trajectories formed by the time-varying values and the model-predicted adverse outcome risks. More specifically, two derived variables the temporal mean and the temporal slope -- are defined, and the associations of the two derived variables with the outcome are expressed in terms of impact scores and impacts, which extends the impact scores and impacts defined for tabular data. The temporal mean can be interpreted as the overall level of the characteristic represented by the clinical concept, and the temporal slope is the linear trend of the characteristic over time. For these associations, interpretations in words in the manner of odds ratios are also provided, which makes the results of the XAI method understandable to clinicians who are familiar with logistic regression models. The XAI method is then applied to the study of CRF and its associations with ADRD risks. In this application, an HVAT model is trained using the CRF data as the longitudinal data and demographic data as the non-longitudinal data. The XAI method quantifies the associations of the temporal mean and temporal slope in CRF trajectories with ADRD risks in terms of impact scores and impacts, whose interpretations are also provided.

It is worth noting that the XAI method is designed to be general and hence not limited to the HVAT models: it can also be applied to other types of DNN models including recurrent neural network (RNN) and its variants such as long short-term memory (LSTM), provided that the input data contains time-varying numerical values similar to those for the HVAT models.

The new XAI method belongs to the class of the perturbation-based XAI methods.^31^ The basic idea of a perturbation-based method is that a black-box model can be better understood through perturbing one input variable at a time and observe how the output value changes along with that. This works straightforward for models taking tabular data as input, because each variable takes only one value on each patient. For HVAT models, an input variable for the longitudinal data is a temporal variable taking multiple values over multiple time points (temporal indices) on a patient, and there are many ways to perturb the multiple values, such as perturbing the first value, perturbing the last value, etc. However, not all such perturbations are easily interpretable. For this study, under the consideration of its application to the study of CRF and ADRD, the perturbation is chosen to be exerted on two derived variables --temporal mean and temporal slope. The challenge is that, only one derived variable can be changed at a time, and the change must be achieved through changing the original sequence of values. This challenge is addressed by introducing two operations named A and B, which perturb the two derived variables separately.

The consideration for the two derived variables is both clinical and mathematical. Clinically, there is an association of the overall CRF level with the ADRD risk and the overall CRF level can be represented by temporal mean of CRF. As the patient ages, the CRF level deteriorates naturally. For two patients with the same overall CRF level but different speed of deterioration, their ADRD risks are also likely different. We hypothesize that if a patient maintains or improves the CRF levels through regular physical activity, the ADRD risk may be lowered. This motivates us to consider both the temporal mean and temporal slope of CRF. This way of characterizing sequence data may be generalized to other clinical variables if the same clinical consideration applies to them. Mathematically, the temporal mean *α* and the temporal slope *β* are exactly the two parameters in the linear equation 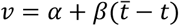 for the line fitted to the sequence of CRF values. To characterize a CRF sequence that is approximately linear in time, these two parameters are natural to consider.

The result on the impact score of temporal mean of CRF shows that higher overall CRF levels are generally associated with lower ADRD risks, and the result on the impact score of the temporal slope of CRF shows that faster deteriorating CRF are generally associated with higher ADRD risks. Both results are consistent with the literature.^32-34^ The previously unknown findings include the following: 1) For patients whose CRF is stable or even increasing over time, a one-unit increase in the speed of increasing CRF has nearly zero effect on the ADRD risks, which is different for those with deteriorating CRF. 2) For patients with lower overall CRF levels, a one-MET increase in the overall CRF level is associated with smaller reduction in ADRD risks than for those with higher overall CRF levels. Both of these discoveries are typical non-linear effects, which demonstrates the advantage of having a DNN model (such as HVAT) over a linear logistic regression model. These discoveries also show that the HVAT model has successfully learned some temporal patterns relevant to the outcome prediction from the data, without explicit human instruction through feature engineering (e.g., explicitly using the temporal mean and slope as input variables), and that the XAI method is effective in revealing the potentially non-linear associations between the temporal patterns and outcome risks.

### Limitations and future work

The XAI method introduced in this paper is an initial attempt to explain a Transformer-based DNN model for clinical outcome prediction. Therefore, we have taken a simple approach, which constitutes the main limitations of the paper. More specifically, limitations include the following.

First, the XAI method is only applicable to the part of the longitudinal input data which are clinical concepts with associated numerical values. There are also clinical concepts without any numerical values such as diagnostic codes, for whose temporal patterns new explanations are needed.

Second, even for the clinical concepts with associated numerical values, there are at least two distinct types. The first type of such concepts are the measurement results of patient health status such as the CRF in this study. Other examples include lab test orders with associated lab values, and vital signs with associated vital sign values. The second type of those are the interventions with strength values. Typical examples of such concepts include drugs with total dosages from prescriptions or refills as the associated values. The multiple values (over multiple time points) associated with such a concept often have a “cumulative” effect on the outcome, making the temporal mean and temporal slope not clinically meaningful on these values, hence the XAI method is not appropriate for this type of concepts. New clinically meaningful derived variables need to be defined for the second type.

Third, we only considered in this study the explanation in terms of linear temporal patterns, which are sufficiently characterized by the temporal mean and temporal slope. There exist nonlinear temporal patterns such as accelerated increase or decrease, periodic oscillations, etc., for which additional derived variables need to be defined.

For the future work, we plan to address all the above limitations.

## Data Availability

Population-level aggregated data in this study are available upon reasonable request to the authors

## Ethics Approval

This study has received an Institutional Review Board (IRB) exemption #1576748-1 from the Washington DC VA IRB.

## Funding

This study was funded by grants NIH/NIA RF1AG069121 and NIH/NIA 1R01AG073474-01A1.

## Notes

### Competing Interest Statement

The authors have declared no competing interest.

### Author Declarations

IRB of Washington DC VA gave ethical approval for this work.

